# DIMR Score: A Tool for Determining the Destination of LVO Patients After Thrombolysis

**DOI:** 10.1101/2023.09.08.23295289

**Authors:** Rahul Rao, Aizaz Ali, Zeinab Zoghi, Julie Shawver, Richard Burgess, Syed Zaidi, Mouhammad Jumaa

## Abstract

**Background:** Stroke patients with large vessel occlusion (LVO) benefit from thrombolysis (tPA) and mechanical thrombectomy (MT). We aim to characterize triaging patterns in these patients, specifically those who go to perfusion-based imaging first or direct to angio in the drip-and-ship model. Furthermore, we propose that select patients may benefit from CTP prior to MT.

**Methods:** A total of 270 patients with acute ischemic stroke secondary to LVO/MeVO were retrospectively evaluated from January 2018 to June 2022. These patients received intravenous tPA from the outside hospital and were transferred for the intention of MT. We compared baseline characteristics between those who received CTP upon arrival and those who went either directly to the angiography suite (DTA) along with logistic regression and functional outcomes.

**Results:** Predictors of CTP utilization over DTA was the presence of an M3 occlusion (11.3% vs. 1.7%, p=0.005) and PCA occlusion (12.9% vs. 3.4%, p=0.015). The DTA approach was higher in M1 MCA occlusions (43.2% vs. 27.4%, p=0.038) and basilar occlusions (7.6% vs. 0, p=0.026). DTA patients had a higher NIHSS at the spoke (median NIHSS 15 [9-21] vs. 9 [4.75-14], p<0.001) and the hub (14 [7-20] vs. 7 [3-15.75], p<0.001). There was no significant difference between the DTA and CTP groups in regards to mRS at 90 days (39% vs. 48.4%, p=0.101).

**Conclusion:** In the drip-and-ship model, NIHSS and location of an occlusion on initial CTA guide CTP utilization in LVO/ MeVO patients. Long term functional outcomes are not significantly affected by arrival at CTP over DTA. Patients whose transfer is delayed, improve after thrombolysis, present with a MeVO, or are limited by resources at the CSC may benefit from transfer to CT over a DTA approach. We propose this DIMR score may help guide triaging of patients who have an intracranial occlusion and receive thrombolysis.

## Introduction

Mechanical thrombectomy (MT) for acute stroke has emerged as an important intervention to improve functional independence. Advanced imaging techniques such as computerized tomography perfusion (CTP) allow appropriate selection of these patients in the late time window^1,2^. In this late window, defined as beyond 6 hours from last seen well, patients are ineligible for thrombolysis (IV tPA or TNK) in the U.S., but those who have an anterior circulation large vessel occlusion (LVO) and slow infarct growth rates as defined by CTP benefit from MT.

In the early time window, specifically in those patients who are receive alteplase (tPA), advanced imaging is thought to be unnecessary when triaging LVO patients for MT^3,4^. Indeed, there are trials that support direct transfer of these patients to the angio suite (DTA) without additional imaging, especially in the early time window^5,6^. However, two MT trials did use CTP in the 6 hour window and showed that target mismatch between ischemic core and penumbra helped to identify those LVO patients who would benefit from thrombectomy^7,8^.

We aim to characterize ischemic stroke patients with LVO who have received thrombolytics and were transferred to our comprehensive stroke center which relies primarily on a DTA approach. Furthermore, we propose that advanced perfusion-based imaging at the CSC may be useful in well selected patients for MT who have received IV thrombolysis at a spoke hospital (drip-and-ship model)^9,10^ without necessarily affecting long-term outcomes.

## Methods

With institutional review board approval, two hundred and seventy patients at a large comprehensive stroke center (CSC) were retrospectively evaluated from 2018-mid 2022. We selected those patients who received IV tPA at a spoke hospital and were transferred to the CSC for the intention of mechanical thrombectomy. Of those, 198 patients had confirmed LVO on CTA. Our practice uses CT scan to triage acute stroke interventions. After the initial non-contrast CT head and CT angiography (CTA) at an outside hospital or spoke, transferred patient with suspected or confirmed LVO or medium vessel occlusion (MeVO), arrive at one of 3 locations: the angio suite directly, the CT scanner at the CSC for additional, usually perfusion-based imaging, or direct admission to our neurological intensive care unit (NICU) if complete resolution of symptoms or medical complications warranting neurocritical care is documented. We separated out those patients who received follow-up imaging, specifically CTP upon arrival, those who went direct to the angiography suite and had groin puncture, those who went direct to the angiography suite and did not have groin puncture, and finally those who were transferred to the NICU (Figure 1). We also separated out patients who improved on the National Institute of Health Stroke Scale (NIHSS) between the spoke and hub and those who worsened.

**Figure 1.**
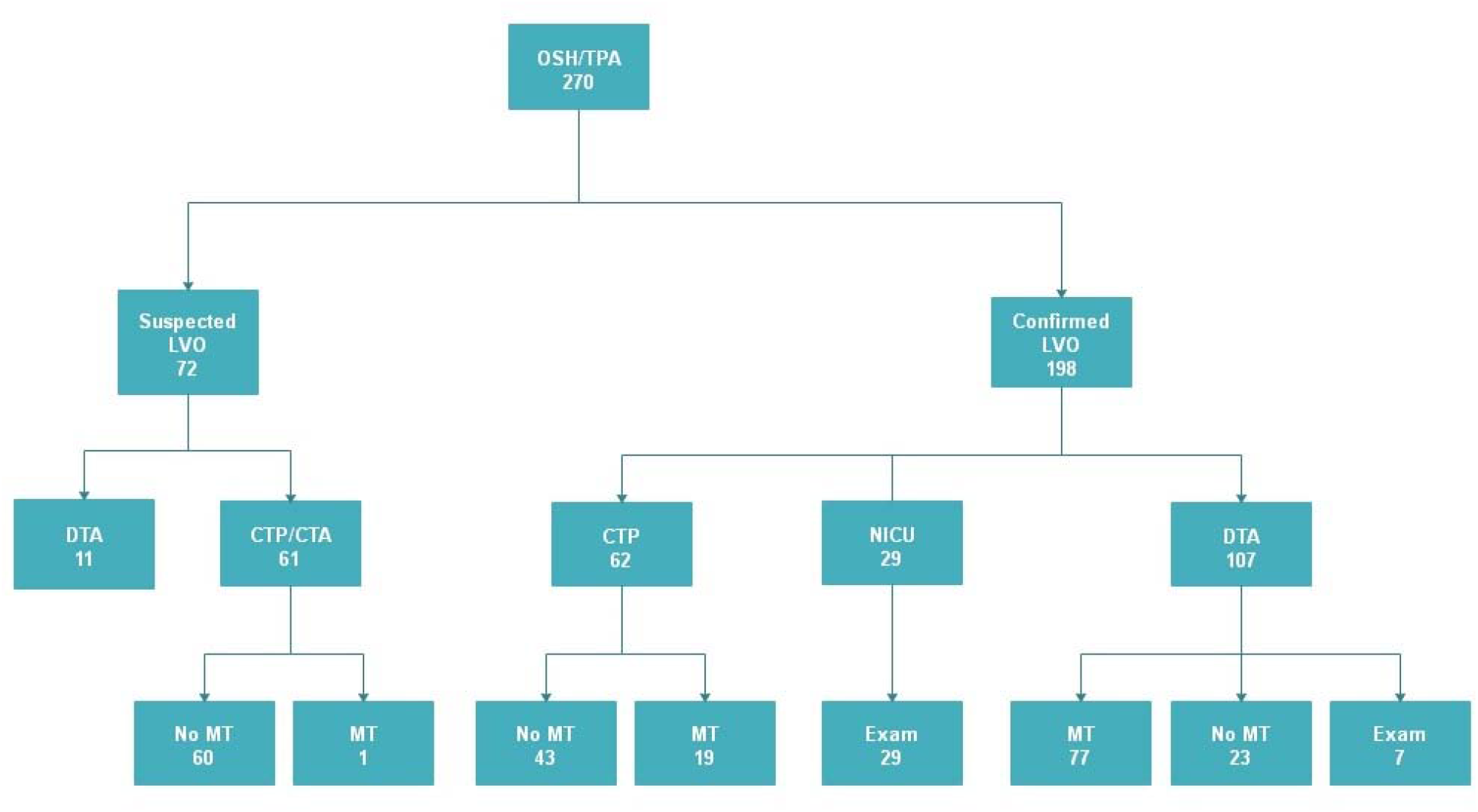
Triaging of LVO Stroke Patients Receiving IV tPA

Baseline demographic variables were obtained from all patients. Our primary time metric was door to groin puncture time and the primary efficacy metric was percentage of favorable outcome defined as modified Rankin Scale (mRS) 0-2 as measured at 90 days. We opted to use groin puncture as opposed to first pass as the primary time metric as some patients recanalized after thrombolysis. We also measured angiographic complications and categorized these as groin (hematoma, femoral thrombosis, etc.) or intracranial (only if clearly iatrogenic from procedure) as our primary safety outcome.

Secondary outcomes included spoke door-to-needle (DTN) time, stroke onset/last seen well to outside hospital arrival, onset to hub arrival, onset to groin puncture time, onset to recanalization time in those receiving thrombectomy, door at hub to perfusion time in those receiving CTP, door to recanalization, first pass effect, intra-arterial thrombolysis use, TIMI/TICI recanalization scores, procedural complications, hospital discharge NIHSS and mRS, discharge disposition, rate of hemorrhagic conversion, mRS and death at 90 days.

We excluded patients who underwent CTP prior to thrombolysis or at the spoke, patients who received CT scan and IV tPA in a mobile stroke unit, and patients who were transferred after thrombolysis directly to MRI as these patients generally do not have a concern for a large vessel occlusion in our practice. We defined resource limitation as times when patients were transferred during the COVID pandemic or when there were concurrent procedures occupying the angio suite such as another mechanical thrombectomy.

### Statistical analysis

Patient were divided into two groups. Group one (CTP) included patients with confirmed LVO on CTA at a referring hospital who went to CTP upon arrival at the CSC. Group two (DTA) included patients who went directly to angio with either suspected or confirmed LVO or MeVO. The univariate comparisons were performed using chi-square test for dichotomous variables and the two sample T-test or Mann-Whitney U test for continuous variables. The normality was calculated using Shapiro-wilk test in the latter variables in order to present and compare them with either mean or median. Multivariable logistic regression was carried out to classify the patients into CTP and DTA after adjustment for baseline, previous history, imaging site of occlusion, procedural and time metric variables. The effect size was measured using Cohen’s d test suggesting that d=0.2, d=0.5, and d=0.8 represent the small, medium, and large group differences, respectively^11^.

The covariates of the baseline patients’ characteristics were balanced across the exposed (the patients who were sent to CTP) and unexposed (those who were directly sent to angio) groups using Inverse Probability Treatment Weighting (IPTW). The effect of CTP as a treatment approach on the favorable outcome (mRS in 90 days <=2) was computed using the logistic regression adjusted with the calculated propensity scores considering the baseline characteristics that are statistically significant.

The univariate analyses were repeated on the patients categorized into those whose NIHSS were improved and not improved to compare the prespecified variables in each group.

The analyses were conducted using SPSS, version 27. The hypothesis testing was made using 2-sided statistical tests with the ≤0.05 significance level.

## Results

Comparing the DTA and CTP groups (Table 1), baseline demographics were significantly different in regards to a higher proportion of coronary artery disease (33.1% vs. 1.1%, p=0.015). The DTA approach was employed more often with M1 occlusion (43.2% vs. 27.4%, p=0.038). Conversely, the CTP group had a higher proportion of M3 occlusion compared to DTA (11.3% vs. 1.7%, p=0.005). Additionally, there were more patients with basilar occlusion in the DTA group (7.6% vs. 0, p=0.026), and more patients with PCA occlusion in the CTP group (12.9% vs. 3.4%, p=0.015). More characteristics were also analyzed as seen in appendix A.

**Table 1.** Baseline characteristics between those who present to CT perfusion (CTP) vs. directly to angiography (DTA)

| Factors | CTP (N=62) | DTA (N=118) | P-Value | LR P-Value |
| --- | --- | --- | --- | --- |
| AGE | 74 [60-81.25] | 73 [61-83.25] | 0.972 | 0.713 |
| GENDER |  |  | 0.079 | 0.081 |
| Male | 22 (35.5%) | 58 (49.2%) |  |  |
| Female | 40 (64.5%) | 60 (50.8%) |  |  |
| HTN | 52 (83.9%) | 103 (87.3%) | 0.529 | 0.530 |
| DM | 21 (33.9%) | 43 (36.4%) | 0.732 | 0.732 |
| HBA1C | 6.37±1.79 | 6.40±1.71 | 0.912 | 0.910 |
| AFIB | 22 (35.5%) | 53 (44.9%) | 0.223 | 0.224 |
| DYSLIPIDEMIA | 44 (71%) | 82 (69.5%) | 0.837 | 0.837 |
| LDL | 84 [63.75-117] | 75 [62-111] | 0.300 | 0.475 |
| TG | 103.5 [82.75-136.5] | 103.5 [76.75-158] | 0.820 | 0.238 |
| HDL | 41 [32-52.25] | 40.5 [33-46] | 0.432 | 0.195 |
| SMOKING | 23 (37.1%) | 39 (33.1%) | 0.587 | 0.587 |
| CAD | 10 (1.1%) | 39 (33.1%) | 0.015 | 0.018 |
| OSH NIHSS | 9 [4.75-14] | 15 [9-21] | <0.001 | <0.001 |
| CSC NIHSS | 7 [3-15.75] | 14 [7-20] | <0.001 | 0.002 |
| ASPECT at OSH | 10 [9-10] | 10 [9-10] | 0.753 | 0.482 |
| ONSET TO OSH ARRIVAL (MINS) | 59.5 [30-110.5] | 49 [30-94] | 0.501 | 0.287 |
| ONSET TO CSC ARRIVAL (MIN) | 242 [184.5-291.5] | 195 [153.5-241.5] | 0.001 | 0.044 |
| CSC DOOR TO GROIN PUNCTURE (MIN) | 59.5 [37-78.75] | 47.25 [18.75-42.5] | <0.001 | 0.407 |
| ONSET TO GROIN PUNCTURE (MIN) | 282.5 [227-326.25] | 235 [181.5-290.5] | 0.047 | 0.382 |
| ONSET TO RECANALIZATION (MIN) | 304 [267-363] | 264 [210.25-337.25] | 0.099 | 0.325 |
| CSC DOOR TO RECAN (MIN) | 90 [71-122] | 57 [44-90] | 0.021 | 0.125 |
| PUNCTURE TO RECAN (MIN) | 24 [15-41] | 27 [17-48.5] | 0.523 | 0.371 |
| Door to CTP (min) | 17 [12.5-26] | 32 [16-32] | 0.217 | 0.571 |
| DTN time | 61 [47.75-80.75] | 50 [38-72.25] | 0.011 | 0.084 |
| MCA M1 | 17 (27.4%) | 51 (43.2%) | 0.038 | 0.039 |
| MCA M2 | 20 (32.3%) | 29 (24.6%) | 0.271 | 0.272 |
| M3 | 7 (11.3%) | 2 (1.7%) | 0.005 | 0.015 |
| M4 | 0 | 1 (0.8%) | 0.467 | 1 |
| ICA T | 3 (4.8%) | 14 (11.9%) | 0.126 | 0.138 |
| PROXIMAL ICA | 8 (12.9%) | 10 (8.5%) | 0.347 | 0.350 |
| TANDEM | 4 (6.5%) | 12 (10.2%) | 0.405 | 0.409 |
| BASILAR | 0 | 9 (7.6%) | 0.026 | 0.999 |
| PCA | 8 (12.9%) | 4 (3.4%) | 0.015 | 0.023 |
| V.A | 2 (3.2%) | 3 (2.5%) | 0.791 | 0.791 |
| SIDE |  |  | 0.395 | 0.186 |
| Posterior | 3 (4.8%) | 11 (9.3%) |  |  |
| Left | 33 (53.2%) | 67 (56.8%) |  |  |
| Right | 26 (41.9%) | 40 (33.9%) |  |  |
| OSH CTA LVO | 60 (96.8%) | 107 (90.7%) | 0.133 | 0.152 |
| IA TPA | 4 (6.5%) | 7 (5.9%) | 0.890 | 0.890 |
| Transport |  |  | 0.381 | 0.906 |
| Day/Night Arrival |  |  | 0.071 | 0.073 |
| Night | 21 (33.9%) | 26 (22%) |  |  |
| Day | 35 (56.5%) | 82 (69.5%) |  |  |
| Angio Complication |  |  | 0.839 | 0.968 |
| None | 16 (25.8%) | 98 (83.1%) |  |  |
| Groin | 0 | 4 (3.4%) |  |  |
| Intracranial | 1 (1.6%) | 4 (3.4%) |  |  |

DTA was associated with higher NIHSS both at the spoke (median NIHSS 15 [9-21] vs. 9 [4.75-14], p<0.001) and the hub (14 [7-20] vs. 7 [3-15.75], p<0.001). Of those receiving thrombectomy, there was a significant difference in median door to groin time between the DTA vs. CTP group (median in minutes [IQR] 47.25 [18.75-42.5] vs. 59.5 [37-78.75], p<0.001) which was our primary time metric.

For our primary efficacy metric, percentage of favorable outcome (mRS 0-2 at 90 days) was similar between the DTA and CTP groups (39% vs. 48.4%, p=0.101). When propensity score analysis was performed, the difference remained statistically insignificant (Table 2). This table represents the results obtained from the logistic regression adjusted with the propensity scores. The logistic regression was adjusted with the statistically significant baseline characteristics as we considered these as potential confounders.

**Table 2.** mRS in 90 days comparison using the logistic regression adjusted with the IPTW weights where the patients were sent to CTP (CTP=1)

| Treatment Approach<br>Favorable Outcome | CTP<br>(N=62) | DTA<br>(N=118) | p-value | OR | 95% of CI |  |
| --- | --- | --- | --- | --- | --- | --- |
|  |  |  |  |  | Lower | Upper |
| mRS [0-2] | 30 (48.4%) | 46 (39%) | 0.144 | 0.541 | 0.237 | 1.234 |

Of the 123 confirmed or suspected LVO patients who received a CT upon arrival to the CSC, 3 received groin puncture only and 18 received MT. Of the 118 DTA patients, 26 received groin puncture only and 82 received MT. DTA was associated with higher odds of groin puncture (OR: 52.5, p<.00001) and MT (OR: 13.3, p<.00001).

For our primary safety outcome, 151 patients total had groin puncture, with 4 having groin complications and 5 having intracranial complications. This was not statistically significant between the DTA and CTP groups (6.8% vs. 1.6%, p=0.839). Of the groin complications, one patient’s angiography revealed tPA had lysed the clot, but the procedure was complicated by popliteal artery occlusion. She underwent popliteal thrombectomy, femoral artery endarterectomy and 4 compartment fasciotomy. NIH had gone from 5 to prior to 0 prior to angio. In another patient, the NIHSS had improved by 12 points but angiography was still performed showing chronic changes so no thrombectomy was performed. This procedure was complicated by a small groin hematoma with ecchymosis. Two other patients had proximal anterior circulation LVO without much improvement prior to thrombectomy and they suffered minor groin hematomas. Of the intracranial complications, one patient had an iatrogenic microwire M2 perforation which was self-resolving, another has an iatrogenic carotid-cavernous fistula after a stent retriever pass and ultimately was transitioned to comfort care after failed thrombectomy. Another patient had an ICA/MCA tandem occlusion complicated by carotid stent thrombosis, but ultimately this patient achieved an mRS of 2. Two patients with left M2 occlusion also had intracranial complications, one with an M3 perforation necessitating coil embolization and another one had a large parenchymal hemorrhage after reperfusion injury following an aspiration pass. This patient was transitioned to comfort care and passed away in-house.

Significant findings amongst our secondary outcomes included the DTA group having a shorter symptom onset to CSC arrival time (195 [153.5-241.5] vs. 242 [184.5-291.5], p=0.001) compared to the CTP group. Time of onset to groin time was also lower in the DTA group (282.5 [227-326.25] vs. 235 [181.5-290.5], p<0.047). The DTA group had a significantly higher proportion of hemorrhagic transformation compared to CTP (31.4% vs. 16.1%, p=0.025), but this was not seen with parenchymal hematoma (4.2% vs. 1.6%, p=0.347) or mortality (17.8% vs. 12.9%, p=0.516).

The distribution of mRS at 90 days between the DTA and CTP groups was also similar (Figure 2) when omitting those patients with missing values at that time.

**Fig 2.**
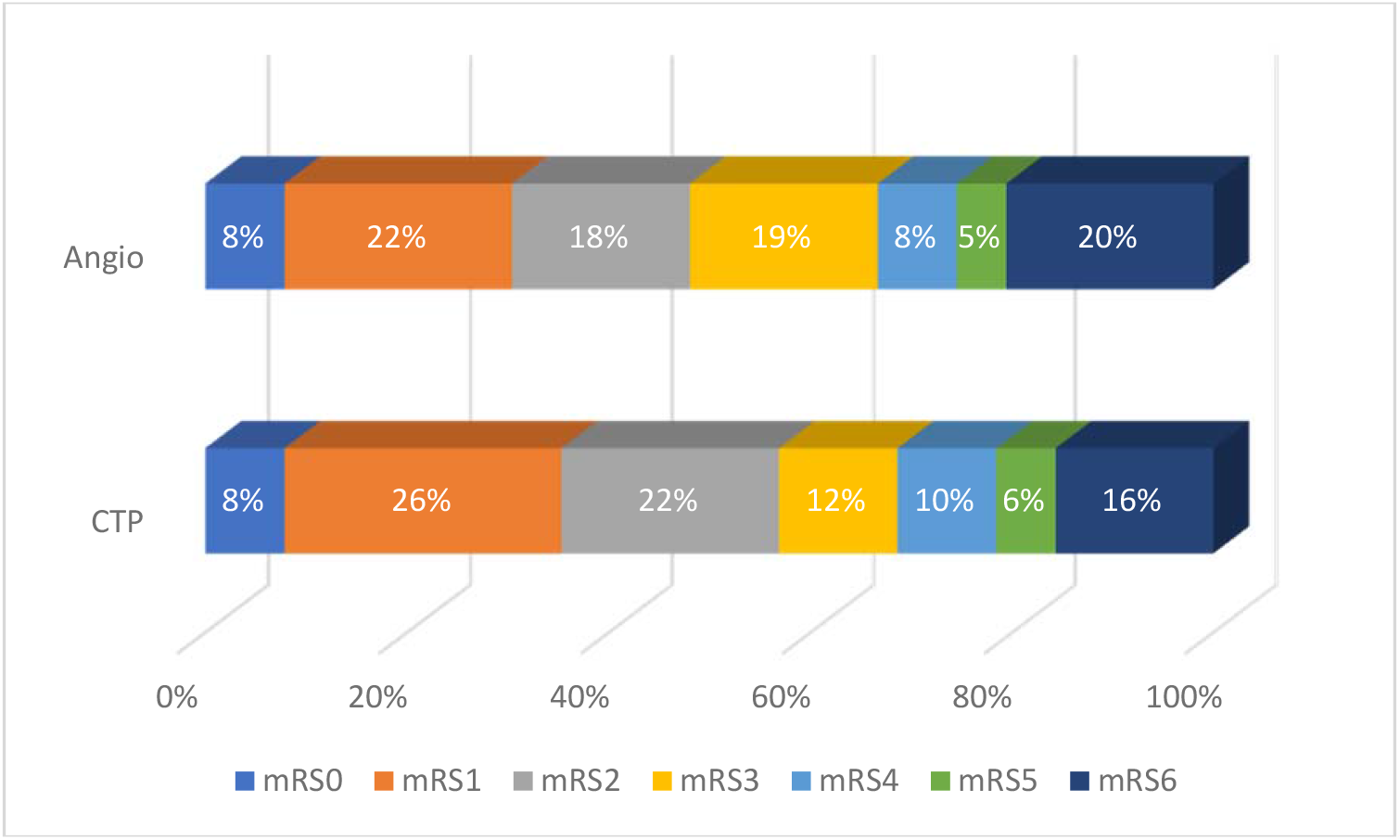
Modified Rankin Scale Plot of patients presenting DTA vs. CTP

When comparing patients whose exam improved and those who worsened (Table 3), CTP utilization was higher in those patients whose NIHSS worsened (49.3% vs. 34.1%, p=0.034). Unsurprisingly, those whose NIHSS worsened had lower rates of favorable outcomes (42.3% vs. 53%, p=0.007) and higher rates of death (23.9 vs. 6.1%, p=0.001) at 90 days.

**Table 3.** Baseline characteristics between those who worsen in transit (Δ NIHSS>0) and those who improve (Δ NIHSS<0)

| Factors | ΔNIHSS>0 (N=71) | ΔNIHSS<0 (N=132) | P-Value |
| --- | --- | --- | --- |
| AGE | 72 [60-81] | 77 [64-84] | 0.095 |
| GENDER |  |  | 0.652 |
| Male | 31 (43.7%) | 62 (47%) |  |
| Female | 40 (56.3%) | 70 (53%) |  |
| HTN | 57 (80.3%) | 104 (78.8%) | 0.802 |
| DM | 25 (35.2%) | 55 (41.7%) | 0.369 |
| HBA1C | 6.39±1.73 | 5.9 [5.5-6.7] | 0.737 |
| AFIB | 26 (36.6%) | 39 (29.5%) | 0.303 |
| DYSLIPIDEMIA | 41 (57.7%) | 93 (70.5%) | 0.068 |
| LDL | 94 [68-122] | 77.5 [63.75-116.25] | 0.079 |
| TG | 109 [77-155] | 98.5 [74.5-140.25] | 0.292 |
| HDL | 42 [32-47] | 40 [34.75-51] | 0.430 |
| SMOKING | 26 (36.6%) | 44 (33.3%) | 0.639 |
| CAD | 14 (19.7%) | 37 (28%) | 0.193 |
| OSH NIHSS | 10 [4-14] | 10 [6-18] | 0.176 |
| TTH NIHSS | 15 [7-19.5] | 4 [2-8.75] | <0.001 |
| ASPECT | 10 [9-10] | 10 [9-10] | 0.135 |
| ONSET TO OSH ARRIVAL (MIN) | 46 [29.5-77.75] | 55 [30-105] | 0.155 |
| ONSET TO ONSITE ARRIVAL (MIN) | 201 [157-280] | 218 [165-290] | 0.220 |
| DOOR TO GROIN PUNCTURE (MIN) | 34 [24.25-76.25] | 28.5 [20-46] | 0.092 |
| ONSET TO GROIN PUNCTURE (MIN) | 230 [172-305.25] | 229.5 [187.5-282.25] | 0.952 |
| ONSET TO RECANALIZATION (MIN) | 266.5 [218.25-349.5] | 274 [189.75-336.25] | 0.501 |
| DOOR TO RECAN (MIN) | 77 [53.75-117] | 65 [44.5-95.5] | 0.205 |
| PUNCTURE TO RECAN (MIN) | 27 [17.25-46.25] | 26 [15.5-55.5] | 0.847 |
| Door to CTP (MIN) | 19 [12-49.75] | 15 [8-26] | 0.292 |
| DTN time | 58 [42-85] | 55 [44.25-76] | 0.767 |
| MCA M1 | 21 (29.6%) | 29 (22%) | 0.242 |
| MCA M2 | 18 (25.4%) | 30 (22.7%) | 0.696 |
| M3 | 5 (7%) | 3 (2.3%) | 0.098 |
| M4 | 0 | 1 (0.8%) | 0.460 |
| ICA T | 4 (5.6%) | 7 (5.3%) | 0.931 |
| PROXIMAL ICA | 7 (9.9%) | 14 (10.6%) | 0.854 |
| TANDEM | 4 (5.6%) | 6 (4.5%) | 0.742 |
| BASILAR | 3 (4.2%) | 7 (5.3%) | 0.727 |
| PCA | 5 (7%) | 12 (9.1%) | 0.605 |
| V.A | 2 (2.8%) | 7 (5.3%) | 0.406 |
| SIDE |  |  | 0.087 |
| Posterior | 5 (7%) | 24 (18.2%) |  |
| Left | 40 (56.3%) | 69 (52.3%) |  |
| Right | 26 (36.6%) | 39 (29.5%) |  |
| OSH CTA LVO | 58 (81.7%) | 94 (71.2%) | 0.101 |
| IV TPA | 71 (100%) | 131 (99.2%) | 0.462 |
| IA TPA | 5 (7%) | 3 (2.3%) | 0.194 |
| TIMI RECANALIZATION |  |  | 0.429 |
| 0 | 3 (4.2%) | 3 (2.3%) |  |
| 1 | 1 (1.4%) | 4 (3%) |  |
| 2 | 26 (36.6%) | 37 (28%) |  |
| 3 | 41 (57.7%) | 88 (66.7%) |  |
| TICI RECANALIZATION |  |  | 0.512 |
| 0 | 3 (4.2%) | 3 (2.3%) |  |
| 1 | 1 (1.4%) | 4 (3%) |  |
| 2 |  | 32 (24.2%) |  |
| 3 | 21 (29.6%) | 88 (66.7%) |  |
| 4 | 40 (56.3%) |  |  |
| 2A |  |  |  |
| 2C |  |  |  |
| TICI 2B+ | 65 (91.5%) | 124 (93.9%) | 0.522 |
| HOSPITAL DC NIHSS | 5.5 [2-11] | 1 [0-4] | <0.001 |
| HOSPITAL DC MRS | 4 [3-5] | 2 [1-4] | <0.001 |
| Change in NIHSS | 4 [2-7] | -4 [-8 – -2] |  |
| DC Disposition |  |  | 0.008 |
| H | 25 (35.2%) | 76 (57.6%) |  |
| IPR | 18 (25.4%) | 27 (20.5%) |  |
| SNF | 15 (21.1%) | 21 (15.9%) |  |
| LTAC | 1 (1.4%) | 3 (2.3%) |  |
| Hos | 6 (8.5%) | 3 (2.3%) |  |
| Dec | 6 (8.5%) | 2 (1.5%) |  |
| 3 MO MRS | 3 [1-6] | 2 [0-3] | <0.001 |
| Favorable Outcome (MRS0-2) | 30 (42.3%) | 70 (53%) | 0.007 |
| DEAD AT 3 MO | 17 (23.9%) | 8 (6.1%) | 0.001 |
| COMPLICATION ICH (HT) | 18 (25.4%) | 24 (18.2%) | 0.240 |
| COMPLICATION ICH (PH) | 2 (2.8%) | 3 (2.3%) | 0.818 |
| COMPLICATION (DISSECTION) | 1 (1.4%) |  | 0.173 |
| CT in house | 40 (56.3%) | 71 (53.8%) | 0.728 |
| FPE | 12 (16.9%) | 9 (6.8%) | 0.730 |
| Transport |  |  | 0.802 |
| Ground | 35 (49.3%) | 67 (50.8%) |  |
| Air | 36 (50.7%) | 64 (48.5%) |  |
| CTP | 35 (49.3%) | 45 (34.1%) | 0.034 |
| CTA Head | 69 (97.2%) | 122 (92.4%) | 0.170 |
| Day/Night Arrival |  |  | 0.058 |
| Night | 27 (38%) | 35 (26.5%) |  |
| Day | 37 (52.1%) | 88 (66.7%) |  |

**Table 4.** Characteristics of Suspected and Confirmed LVO Patients who went to CT upon CSC Arrival.

|  |  |
| --- | --- |
| DIMR Score | N = 123 |
| **Delayed Transport Time (>2 hours)** | 39 (31.7%) |
| **Improvement in Exam (NIHSS)** | 69 (56.1%) |
| **Medium Vessel Occlusion (MeVO)** | 13 (10.6%) |
| **Resource Limitation** | 41 (33.3%) |
| None of the above | 14 (11.4%) |
| Multiple of the above | 39 (31.7%) |

Of the 123 patients with confirmed or suspected LVO who were transferred to CT, 109 of them (88.6%) either had delayed transport by 2 hours or more, improved on NIHSS upon arrival to the hub, had a MeVO, or were transferred during times of resource limitation.

## Discussion

This study aims to characterize triage patterns of drip-and-ship ischemic stroke patients with LVO upon arriving to a CSC that relies primarily on a DTA approach. While studies have emerged supporting basic imaging for MT selection^12^ and the direct to angio model^5,6^, we propose that for patients that receive thrombolysis, advanced imaging may still play a role in certain patients. CTP use did not lead to worsening outcomes at 90 days and were associated with lower mild hemorrhagic transformation rates. In this era of workflow limitations, employing advanced imaging techniques may better select improving patients for thrombectomy in the early time window and may even lead to better outcomes^13^.

Our results show that triaging after thrombolysis in our transferred LVO patients can vary widely. While we are heavy utilizers of the DTA approach for confirmed LVO, factors that can influence alternative destinations include time of day, operator preference, initial location of occlusion, mode of transport, and examination change. Our study demonstrates that a higher overall NIHSS led to a significantly higher utilization of the direct-to-angio approach. However, more than one-third of patients whose examination improved and almost half of patients who worsened received CTP upon arrival to the hub.

Interestingly, post-tPA patients with an M1 occlusion were more likely to be brought directly to angiography whereas patients who had a more distal M3 occlusion were more likely to receive CTP. This finding suggests that at least amongst middle cerebral artery occlusions, a DTA approach is more likely in a proximal clot which is probably less amenable to spontaneous recanalization. In more medium vessel location (MeVO), when thrombolytics are administered and there is time for the drug to work in transit, employing pause before angiography activation may be a reasonable approach. Obtaining a CTP only requires about 40 mL of IV contrast^14^ and is considerably less than a repeat CT angiogram or conventional angiography.

Similarly, patients with a basilar occlusion were more likely to be transferred directly to angio, whereas patients with PCA occlusions were more likely to be transferred to CTP first. With the high morbidity of basilar occlusion, it makes sense to transfer them directly to angio. Despite a recent publication showing trends towards favorability for endovascular management of PCA occlusion^15^, the lack of randomized trial data indicates that patients with these occlusions may benefit from examination and additional imaging prior to thrombectomy as evidenced in our results.

There are many centers that likely employ a DTA approach when transferring LVO patients, even those who may have gotten thrombolysis and improved. Based on our data, we believe that there are situations where directly transferring to angiography may not be the best initial triage. 132 patients seemingly improved based on NIHSS between the spoke and the hub. Out of the 123 patients presenting directly to CTP, only 18 received mechanical thrombectomy and 3 had groin puncture and no intervention. Out of the 118 patients that went directly to angio, 10 did not receive groin puncture secondary to an improved examination on the table, whereas 26 patients received a groin puncture but they had recanalized on subsequent angiography. Indeed, a DTA approach led to higher odds of groin puncture and mechanical thrombectomy in our cohort.

While we see longer door to groin puncture times in our CTP cohort, there is a non-significant difference in terms of 90 day functional outcome. This is similar to 2017 study examining the DTA approach, but stands in contrast to more recent publications^16,17,18^. Of note, Sarraj et al. described increased mortality and safety concerns in the DTA approach if the transfer time exceeded 3 hours^19^.

In our cohort, the DTA group had a higher hemorrhage rate than the CTP group, although only mild hemorrhagic transformation was statistically significant. Moreover, while overall complication rate for thrombectomy is low, our cohort had a 6% complication rate with groin puncture. Delaying thrombectomy for a CTP did not lead to a higher angio complication rate. One of our patients required major vascular surgery intervention following a groin complication secondary to angiography despite improving to an NIHSS of 0 on arrival. Two other patients were transitioned to comfort care following iatrogenic intracranial complications.

Gao et al suggested that the clinical and economic benefits of pre-MT may be increased in the drip-and-ship model^20^. Recently, Colasurdo et al suggested the HALT score as a predictor for LVO recanalization prior to thrombectomy; with one of the predictors being early thrombolysis^21^. By identifying some of these factors before arrival to the hub, we may be able to avoid unnecessary angiography activation in certain situations and transferring first to advanced imaging may be reasonable. CTP utilization leads to longer door-to-groin puncture times, but if the outcomes at 90 days are similar, CTP may help to triage and better select for those patients who will benefit from a procedure that is expensive, requires man power, and is not without risk.

In LVO patients who do not receive thrombolysis and are therefore not given therapy to help them improve, we employ a direct-to-angio approach. In those with persistent disabling deficits, a DTA approach is still absolutely indicated as evidenced by better outcomes in these patients^22^. However, if the examination has improved to the point where deficits are either absent or non-disabling, advanced imaging may provide reassurance that there isn’t a persistent LVO. Conversely, patients who worsen on examination may also benefit from additional imaging to determine if there is hemorrhage secondary to thrombolysis or if a large penumbra still exists. While this latter point is now less relevant due to the recently published large core thrombectomy trials^23,24,25^, those patients whose core had enlarged in transit and do not have a proximal anterior circulation occlusion may not necessarily benefit from DTA. The lack of clear randomized trial data supporting thrombectomy in MeVOs and non-basilar posterior circulation occlusions also should allow for pause and more data in the form of perfusion-based imaging after thrombolysis.

We propose a novel DIMR score as a tool from which to determine which patients may benefit from transfer to CT prior to the angiography suite. The D stands for delayed transfer greater than 2 hours, I for improvement on examination, M for CTA findings suggestive of a MeVO (M3, M4, A2, P2), and R for those transferred in times of resource limitation. 88.9% of our direct to CT patients fit into one of these categories and 31.7% fit into more than one. While DTA should be the default destination, transferring a patient with one of these characteristics to CTP prior to angio may yield more data, reduce unnecessary angio activation and groin puncture rates.

Although all of the IV thrombolysis in our patients were with alteplase, with the advent of tenecteplase (TNK), patients may have even higher pre-endovascular recanalization rates. The EXTEND-IA TNK part 2 trial specifically included patients in rural and regional centers and found 34% of tenecteplase-treated patients had reperfused by the time of arrival at a thrombectomy-capable hospital^26^. TNK-treated patients may also fit well into the CTP first approach, but more data is needed.

While it may seem controversial to utilize CTP in the early time window, one has to keep in mind that the status quo changes with thrombolysis. A substantial improvement in NIHSS does not mean there is an absence of disabling deficits and should not be the sole determinant of employing the CTP first approach. Indeed, there are several clinical trials being performed examining the utility of low NIHSS thrombectomy^27,28.^ Indeed, one study suggests that excellent functional outcome in these patients may be guided by perfusion imaging^29^. If CTP can be used to help to avoid unnecessarily subjecting a patient to angiography and/or avoid calling in the angiography team, there remains utility for this imaging modality in the early window.

### Limitations

Our study has several limitations. While the majority of our patients had LVOs, we did include some MeVOs as we will routinely bring these patients to our angiography suite for possible thrombectomy. We used change in NIHSS used a metric of clinical improvement or worsening. The DTA group did have a higher baseline NIHSS and therefore likely had more severe stroke which may explain the higher rates of hemorrhage and lower odds of functional improvement. We did try to minimize this bias by employing propensity weighting. Finally, we did not assess degree of perfusion mismatch in those who received CTP. The degree of target mismatch, in addition to examination, likely plays a large role in whether these patients continue on to the angio suite or not.

## Conclusion

In LVO stroke patients who receive thrombolysis during transfer, NIHSS and location of an occlusion on initial CTA guide CTP utilization in a primarily direct-to-angio practice. Utilization of CTP in the early time window leads to delays in groin puncture, but not necessarily functional outcomes. Delayed transfer, improvement in examination, medium vessel occlusion, and resource or workflow limitation may justify CTP utilization in these patients. Future studies are indicated to determine what other parameters may better select the destination for drip-and-ship patients, and if TNK, PCA occlusion, and low NIHSS patients still benefit from a DTA approach, especially after thrombolysis.

## Data Availability

All data supporting the findings of this study is available upon reasonable request to the primary author

## Abbreviations

MT: mechanical thrombectomy
CTP: computerized tomography perfusion
TPA: tissue plasminogen activator/alteplase
DTA: direct to angiography
CTA: computerized tomography angiography
LVO: large vessel occlusion
ICU: intensive care unit
CSC: comprehensive stroke center
MRI: magnetic resonance imaging
NIHSS: National Institute of Health Stroke Scale
TNK: tenecteplase
DTN: door-to-needle
TIMI: thrombolysis in myocardial infarction
TICI: thrombolysis in cerebral infarction
mRS: modified Rankin Scale
(MeVO): medium vessel occlusions
DIMR: delayed, improved, MeVO, resource limitation,

## Acknowledgments

None

## Appendix A

| Factors | CTP (N=62) | DTA (N=118) | P-Value |
| --- | --- | --- | --- |
| TIMI RECANALIZATION |  |  | 0.110 |
| 0 | 3 (4.8%) | 3 (2.5%) |  |
| 1 | 4 (6.5%) | 1 (0.8%) |  |
| 2 | 25 (40.3%) | 45 (38.1%) |  |
| 3 | 30 (48.4%) | 69 (58.5%) |  |
| TICI RECANALIZATION |  |  | 0.079 |
| 0 | 3 (4.8%) | 3 (2.5%) |  |
| 1 | 4 (6.5%) | 0 |  |
| 2 | 0 | 1 (0.8%) |  |
| 3 | 21 (33.9%) | 34 (28.8%) |  |
| 4 | 29 (46.8%) | 66 (55.9%) |  |
| 2A | 1 (1.6%) | 3 (2.5%) |  |
| 2C | 4 (6.4%) | 11 (9.3%) |  |
| TICI 2B+ | 54 (87.1%) | 111 (94.1%) | 0.108 |
| HOSPITAL DC NIHSS | 3 [1-7.75] | 4 [1-9.5] | 0.128 |
| HOSPITAL DC MRS | 3 [2-4] | 4 [2-5] | 0.121 |
| Change in NIHSS | 0 [-3 – 2] | 0 [-4 – 0.25] | 0.453 |
| DC Disposition |  |  | 0.257 |
| H | 30 (48.4%) | 36 (30.5%) |  |
| IPR | 12 (19.4%) | 35 (29.7%) |  |
| SNF | 12 (19.4%) | 27 (22.9%) |  |
| LTAC | 1 (1.6%) | 5 (4.2%) |  |
| Hos | 3 (4.8%) | 8 (6.8%) |  |
| Dec | 4 (6.5%) | 7 (5.9%) |  |
| 3 MO MRS | 2 [1-4] | 3 [1-5] | 0.447 |
| Favorable Outcome (MRS0-2) | 30 (48.4%) | 46 (39%) | 0.101 |
| DEAD AT 3 MO | 8 (12.9%) | 21 (17.8%) | 0.516 |
| COMPLICATION ICH (HT) | 10 (16.1%) | 37 (31.4%) | 0.025 |
| COMPLICATION ICH (PH) | 1 (1.6%) | 5 (4.2%) | 0.347 |
| COMPLICATION (DISSECTION) | 1 (1.6%) | 1 (0.8%) | 0.646 |
| CT in house | 58 (93.5%) | 14 (11.9%) | <0.001 |
| FPE | 6 (9.7%) | 33 (28%) | 0.612 |
| Ground | 29 (46.8%) | 55 (46.6%) |  |
| Air | 32 (51.6%) | 63 (53.4%) |  |
| CTP | 62 (100%) | 9 (7.6%) | <0.001 |
| Night | 21 (33.9%) | 26 (22%) |  |
| Day | 35 (56.5%) | 82 (69.5%) |  |

